# Cost-Effectiveness Analysis of Statins for Treatment of Hospitalized COVID-19 Patients

**DOI:** 10.1101/2021.04.29.21256335

**Authors:** Ronald Chow, Elizabeth Horn Prsic, Hyun Joon Shin

**Affiliations:** Yale School of Public Health, Yale University, New Haven, CT, United States of America; Yale New Haven Health, Yale School of Medicine, Yale University, New Haven, CT, United States of America; Hanyang Impact Science Research Center, Seoul, Korea; Lemuel Shattuck Hospital, Jamaica Plain, MA, United States of America; Brigham and Women’s Hospital, Boston, MA, United States of America

**Author notes:** Correspondence: Ronald Chow, Department of Chronic Disease Epidemiology, Yale School of Public Health, 60 College Street, New Haven, CT 06510, USA, Hyun Joon Shin, Chief of Cardiology, Division of Cardiology, Department of Medicine, Lemuel Shattuck Hospital, Massachusetts, Department of Health, 170 Morton St, Jamaica Plain, MA 02130, USA. Disclosures: None.

**Keywords:** statins, COVID-19, cost-effectiveness analysis

## Abstract

**Introduction:** A recent systematic review and meta-analysis by our group reported on thirteen published cohorts investigating 110,078 patients. Patients administered statins after their COVID-19 diagnosis and hospitalization were found to have a lower risk of mortality. Given this reported superiority, a logical next question would be whether statins are cost-effective treatment options for hospitalized COVID-19 patients. In this paper, we report on a cost-effectiveness analysis of statin-containing treatment regimens for hospitalized COVID-19 patients, from a United States healthcare perspective.

**Methods:** A Markov model was used, to compare statin use and no statin use among hospitalized COVID-19 patients. The cycle length was one week, with a time horizon of 4 weeks. A Monte Carlo microsimulation, with 20,000 samples were used. All analyses were conducted using TreeAge Pro Healthcare Version 2021 R1.1.

**Results:** Treatment of hospitalized COVID-19 patients with statins was both cheaper and more effective than treatment without statins; statin-containing therapy dominates over non-statin therapy.

**Conclusion:** Statin for treatment of COVID-19 should be further investigated in RCTs, especially considering its cost-effective nature. Optimistically and pending the results of future RCTs, statins may also be used broadly for treatment of hospitalized COVID-19 patients.

## INTRODUCTION

The COVID-19 pandemic, as declared by the World Health Organization on March 12, 2020^1^, has been an ongoing pandemic for over a year, at the time of this writing. Many potential treatments have been explored, including statins. Mechanistically, statins may inhibit 3-hydroxy-glutaryl-CoA (HMG-CoA) reductase in cells, and reduce a cytokine storm^2-4^.

A recent systematic review and meta-analysis by our group reported on thirteen published cohorts investigating 110,078 patients. Patients administered statins after their COVID-19 diagnosis and hospitalization were found to have a lower risk of mortality – hazard ratio of 0.53, 95% CI: 0.46-0.61, and odds ratio of 0.57, 95% CI: 0.43-0.75. Given this reported superiority, a logical next question would be whether statins are cost-effective treatment options for hospitalized COVID-19 patients.

In this paper, we report on a cost-effectiveness analysis of statin-containing treatment regimens for hospitalized COVID-19 patients, from a United States healthcare perspective.

## METHODS

### The Model

A Markov model was used, to compare statin use and no statin use among hospitalized COVID-19 patients (Figure 1). There were four assumed health states – “Hospitalized, Non-ICU”, “Hospitalized, ICU”, “Discharged” and “Dead”. Patients treated with statins started in the “Hospitalized, Non-ICU” health state, and may or may not experience a drug-related adverse event during the cycle. If patients experienced liver or muscle toxicity, statins were discontinued.

**Figure 1.**
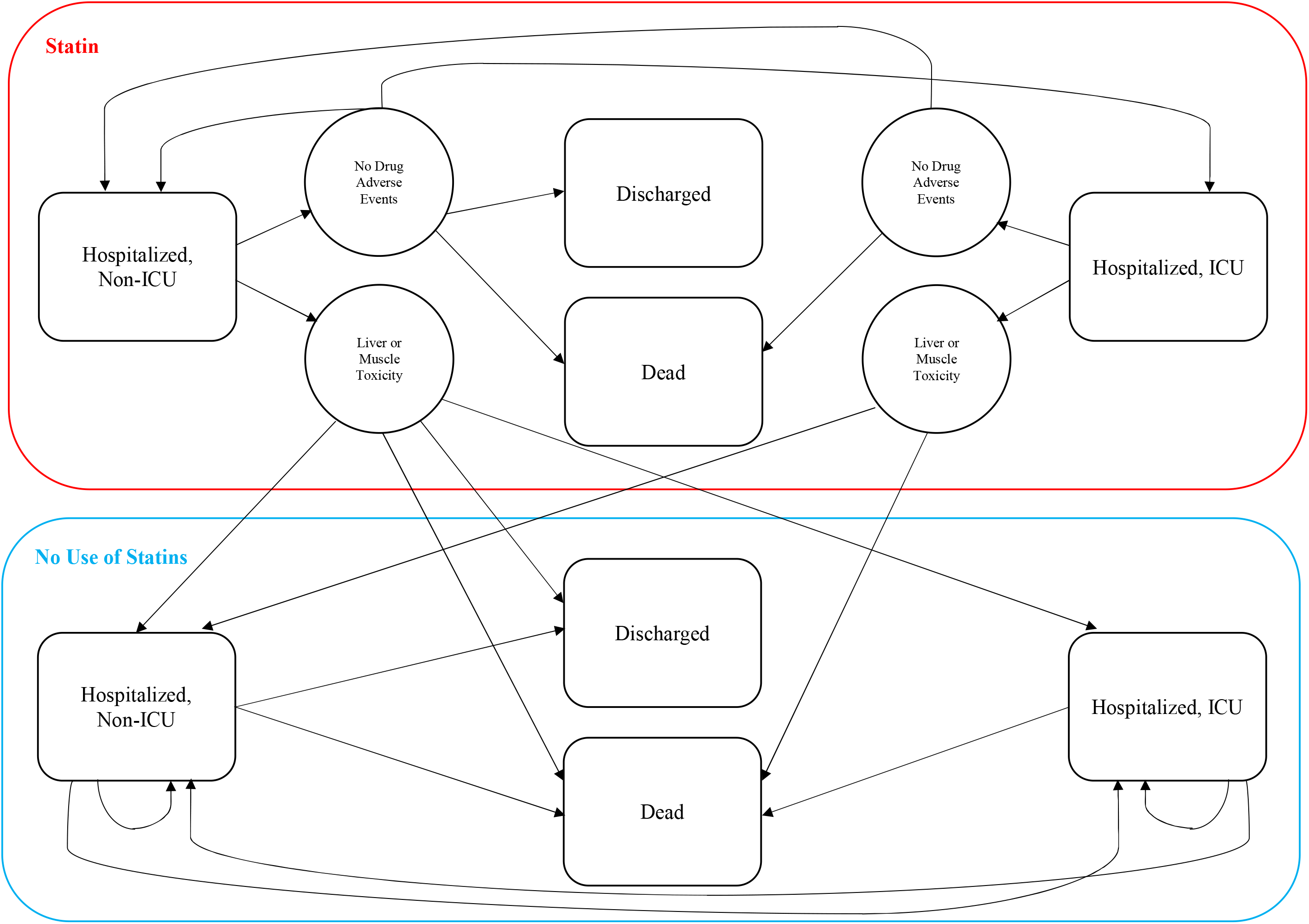
State-Transition Diagram

Patients either remained in the “Hospitalized, Non-ICU” health state, or transitioned to one of the three other health states. For patients who were admitted to the ICU, we assumed that they would be transferred back to non-ICU inpatient care prior to discharge, or died. We also assumed that patients who were discharged would not be readmitted to the hospital for COVID-19, within 1 month.

The cycle length was one week, with a time horizon of 4 weeks. No discounting rate was used, due to the acute timeline. A Monte Carlo microsimulation, with 20,000 samples were used. All analyses were conducted using TreeAge Pro Healthcare Version 2021 R1.1.

### Probabilities

We sourced 1-week probability of death, discharge and ICU admission for non-statin hospitalized COVID-19 patients from Zhang *et al*^5^ and Rodriguez-Nava *et al*^6^. From Zhang *et al*^5^, we computed the 1-week probability from their 4-week Kaplan-Meier curve statistics. We assumed that probabilities reported by Rodriguez-Nava *et al*^6^ were 4-week probabilities, and computed 1-week probability from it. Probability of death among statin patients was computed, using relative risks reported from a prior meta-analysis (Chow et al^7^) and the probability reported by Zhang *et al*^5^ for non-statin patients. Probability of discharge and ICU admissions among statin patients was also sourced from Zhang *et al*^5^; probability of death among ICU patients receiving statins was extracted from Rodriguez-Nava *et al*^6^. The probabilities for liver and muscle toxicity were sourced from Gitlin *et al al*^8^. Beta distributions were used, to model probabilities (Table 1).

**Table 1.**
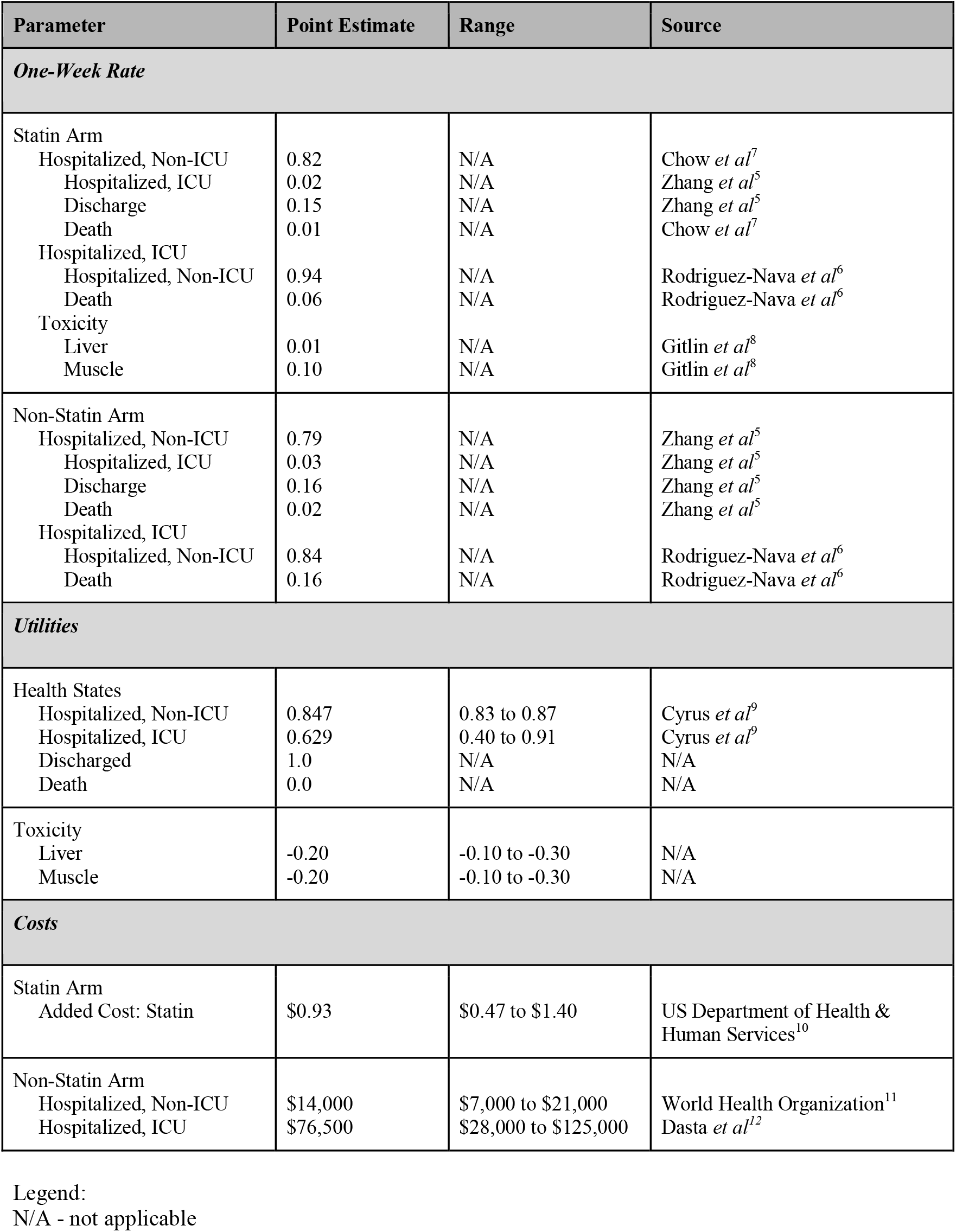
Inputs for Markov Model – Statin Use After Hospitalization

| Parameter | Point Estimate | Range | Source |
| --- | --- | --- | --- |
| **One-Week Rate** |  |  |  |
| Statin Arm |  |  |  |
| Hospitalized, Non-ICU | 0.82 | N/A | Chow *et al*<sup>7</sup> |
| Hospitalized, ICU | 0.02 | N/A | Zhang *et al*<sup>5</sup> |
| Discharge | 0.15 | N/A | Zhang *et al*<sup>5</sup> |
| Death | 0.01 | N/A | Chow *et al*<sup>7</sup> |
| Hospitalized, ICU |  |  |  |
| Hospitalized, Non-ICU | 0.94 | N/A | Rodriguez-Nava *et al*<sup>6</sup> |
| Death | 0.06 | N/A | Rodriguez-Nava *et al*<sup>6</sup> |
| Toxicity |  |  |  |
| Liver | 0.01 | N/A | Gitlin *et al*<sup>8</sup> |
| Muscle | 0.10 | N/A | Gitlin *et al*<sup>8</sup> |
| Non-Statin Arm |  |  |  |
| Hospitalized, Non-ICU | 0.79 | N/A | Zhang *et al*<sup>5</sup> |
| Hospitalized, ICU | 0.03 | N/A | Zhang *et al*<sup>5</sup> |
| Discharge | 0.16 | N/A | Zhang *et al*<sup>5</sup> |
| Death | 0.02 | N/A | Zhang *et al*<sup>5</sup> |
| Hospitalized, ICU |  |  |  |
| Hospitalized, Non-ICU | 0.84 | N/A | Rodriguez-Nava *et al*<sup>6</sup> |
| Death | 0.16 | N/A | Rodriguez-Nava *et al*<sup>6</sup> |
| **Utilities** |  |  |  |
| Health States |  |  |  |
| Hospitalized, Non-ICU | 0.847 | 0.83 to 0.87 | Cyrus *et al*<sup>9</sup> |
| Hospitalized, ICU | 0.629 | 0.40 to 0.91 | Cyrus *et al*<sup>9</sup> |
| Discharged | 1.0 | N/A | N/A |
| Death | 0.0 | N/A | N/A |
| Toxicity |  |  |  |
| Liver | -0.20 | -0.10 to -0.30 | N/A |
| Muscle | -0.20 | -0.10 to -0.30 | N/A |
| **Costs** |  |  |  |
| Statin Arm |  |  |  |
| Added Cost: Statin | \$0.93 | \$0.47 to \$1.40 | US Department of Health & Human Services<sup>10</sup> |
| Non-Statin Arm |  |  |  |
| Hospitalized, Non-ICU | \$14,000 | \$7,000 to \$21,000 | World Health Organization<sup>11</sup> |
| Hospitalized, ICU | \$76,500 | \$28,000 to \$125,000 | Dasta *et al*<sup>12</sup> |

N/A - not applicable

### Effectiveness

Utilities for non-ICU hospitalization and ICU hospitalization were noted from Cyrus *et al*^9^. We assumed utilities for those who were discharged, died, experienced liver toxicity and experienced muscle toxicity. We conservatively assumed a disutility of −0.2 for liver and muscle toxicity, which would be a greater disutility than a patient’s health state changing from non-ICU hospitalization to ICU-hospitalization. Gamma distributions were used, for utility associated with hospitalization. Triangular distributions were used, for disutility associated with liver and muscle toxicity.

### Costs

We calculated the cost of statins from the US Department of Health & Human Services^10^, as the average cost of atorvastatin 10mg, simvastatin 40mg, pravastatin 40mg and lovastatin 40mg. We assumed price to range +/- 50%. The cost for non-ICU hospitalization was calculated using the 1-day average hospitalization cost of approximately $2,000 from the World Health Organization^11^, and varied costs +/- 50%. We used the one-day cost of ICU hospitalization with mechanical ventilation from the Dasta *et al*^*12*^ paper, and calculated a lower-bound range from the one-day cost of ICU hospitalization after stabilization on day 3. The upper-bound was calculated, to produce a symmetric range. A uniform distribution was used, to represent costs.

## RESULTS

The mean cost for patients receiving statins was $31,623 (SD $20,331), whereas the mean cost for patients not receiving statins was $33,218 (SD $25,440). The mean effectiveness for the two cohorts were 1.73 (SD 0.96) and 1.71 (SD 1.00), respectively. Treatment of hospitalized COVID-19 patients with statins was both cheaper and more effective than treatment without statins; statin-containing therapy dominates over non-statin therapy.

## DISCUSSION

To our knowledge, this is the first cost-effectiveness analysis reporting on statins used for the treatment of hospitalized COVID-19 patients. We report herein that treatment with statins, relative to without statins, is both cheaper and more effective.

In an ideal world, this conclusion would carry little weight, as ideally most of the populace would have been vaccinated against COVID-19 and thereby very few incident cases would occur. Unfortunately, distribution of developed vaccines has been slow, and only a few developed countries have enough vaccines for their entire population^13,14^. Furthermore, variants of COVID-19 may escape immunity from vaccination^15^. In light of this, new COVID-19 cases will likely continue to emerge and treatment will be necessary, rather than prevention.

At the time of this writing in April 2021, remdesivir, systemic glucocorticoid and tocilizumab are recommended for patients with severe COVID-19 pneumonia^16^. There is currently no safe treatment for patients with non-severe COVID-19. Given the results of our prior systematic review^7^, and now this cost-effectiveness analysis suggesting statin-containing treatments are both cheaper and more effective, statins should be further investigated for use in non-severe COVID-19 patients. Statin therapy may lead to side effects of myopathy and liver toxicity, but these adverse events typically reverse, after discontinuation of statins.

We eagerly await the results of the ongoing COLSTAT trial, investigating colchicine/statins for the prevention of COVID-19. These results will hopefully provide insight as to whether statins are also appropriate in the prevention setting.

This study was not without limitations. Our underlying probabilities were sourced from observational studies, and therefore patients receiving and not receiving statins may have unbalanced characteristics that may lead to confounding. To account for potential imbalances, we used adjusted probabilities, from multivariable models. We also used beta distributions for probabilities, to account for potential variation in measured effectiveness relative to true effectiveness/efficacy. Another limitation is the omission of other possible side effects inlcuding statin-induced dementia^17^ and statin-induced diabetes^18^. These adverse events are likely of marginal concern in patients with immediate risk of COVID-19 pneumonia, where the interest of treatment is improved health state in a very acute timeline until stabilization after COVID-19 infection. This differs from prior studies and cost-effectiveness analyses of statins in other settings, which report long term side effect of statins, including diabetes and dementia. It is also important to note that the link between dementia and statins is unclear at this time.

In conclusion, treatment of hospitalized COVID-19 patients led to better effectiveness, but also cheaper cost. Statin for treatment of COVID-19 should be further investigated in RCTs, especially considering its cost-effective nature. Optimistically and pending the results of future RCTs, statins may also be used broadly for treatment of hospitalized COVID-19 patients.

## Data Availability

N/A

## REFERENCES

1. Chow R, Elsayed S, Lock M. How robust are the results of one of the first positive trials of hydroxycloroquine for treatment of COVID-19? medRxiv 2020

2. Fedson DS, Opal SM, Rordam OM. Hiding in Plain Sight: an Approach to Treating Patients with Severe COVID-19 Infection. mBio 2020.

3. Scicali R, Di Pino A, Piro S, Rabuazzo AM, Purrello F. May statins and PCSK9 inhibitors be protective from COVID-19 in familial hypercholesterolemia subjects? Nutr Metab Cardiovasc Dis 2020;30:1068–9.

4. Soto-Acosta R, Mosso C, Cervantes-Salazar M, et al. The increase in cholesterol levels at early stages after dengue virus infection correlates with an augment in LDL particle uptake and HMG-CoA reductase activity. Virology 2013;442:132–47.

5. Zhang XJ, Qin JJ, Cheng X, et al. In-Hospital Use of Statins Is Associated with a Reduced Risk of Mortality among Individuals with COVID-19. Cell Metab 2020;32:176-87.e4.

6. Rodriguez-Nava G, Trelles-Garcia DP, Yanez-Bello MA, Chung CW, Trelles-Garcia VP, Friedman HJ. Atorvastatin associated with decreased hazard for death in COVID-19 patients admitted to an ICU: a retrospective cohort study. Crit Care 2020;24:429.

7. Chow R, Im J, Chiu N, et al. The protective association between statins use and adverse outcomes among COVID-19 patients: a systematic review and meta-analysis. medRxiv 2021.

8. Gitlin Z, Marvel F, Blumenthal RS, Martin SS. Statin Safety and Adverse Events. American College of Cardiology 2018.

9. Cyrus A, Safura Y, Farman Zahir A, et al. Health States Utility Value in COVID-19. ResearchSquare 2021.

10. NADAC (National Average Drug Acquisition Cost). Data.gov, 2021. (Accessed 23 April, 2021, at https://catalog.data.gov/dataset/nadac-national-average-drug-acquisition-cost.)

11. ChOosing Interventions that are Cost Effective. World Health Organization, 2020. (Accessed 18 April, 2021, at https://www.who.int/choice/country/usa/cost/en/.)

12. Dasta JF, McLaughlin TP, Mody SH, Piech CT. Daily cost of an intensive care unit day: the contribution of mechanical ventilation. Crit Care Med 2005;33:1266–71.

13. COVID-19 vaccines: resolving deployment challenges. Bull World Health Organ 2021;99:174–5.

14. Burgos RM, Badowski ME, Drwiega E, et al. The race to a COVID-19 vaccine: opportunities and challenges in development and distribution. Drugs Context 2021;10.

15. Hacisuleyman E, Hale C, Saito Y, et al. Vaccine Breakthrough Infections with SARS-CoV-2 Variants. LID - 10.1056/NEJMoa2105000 [doi].

16. Kim AY, Gandhi RT. COVID-19: Management in hospitalized adults. UpToDate 2021.

17. Odden MC, Pletcher MJ, Coxson PG, et al. Cost-effectiveness and population impact of statins for primary prevention in adults aged 75 years or older in the United States. Ann Intern Med 2015;162:533–41.

18. Ganda OP. Statin-induced diabetes: incidence, mechanisms, and implications. F1000Res 2016;5:F1000 Faculty Rev–499.

